# Risk of Sexually Transmitted Infections Among U.S. Military Service Members in the Setting of HIV Pre-Exposure Prophylaxis Use

**DOI:** 10.1101/2022.08.08.22278539

**Authors:** Jason M. Blaylock, Evan C. Ewers, Elizabeth J. Bianchi, David B. King, Rosemary O. Casimier, Hector Erazo, Stephen Grieco, Jenny Lay, Sheila A. Peel, Kayvon Modjarrad, Charmagne G. Beckett, Jason F. Okulicz, Paul T. Scott, Shilpa Hakre

## Abstract

**Background:** The evidence for an increased incidence of sexually transmitted infections (STIs) among patients utilizing HIV pre-exposure prophylaxis (PrEP) has been inconsistent. We assessed the risk of incident STI while on PrEP compared to periods off PrEP among military service members starting PrEP.

**Methods:** Incidence rates of chlamydia, gonorrhea, syphilis, hepatitis C virus, and HIV were determined among military service members without HIV prescribed daily oral tenofovir disoproxil fumarate and emtricitabine for HIV PrEP from February 1, 2014 through June 10, 2016. Hazard ratios for incident STIs were calculated using an Anderson-Gill recurrent event proportional hazard regression model.

**Results:** Among 755 male service members, 477 (63%) were diagnosed with incident STIs (overall incidence 21.4 per 100 person-years). In multivariate analysis, male service members had a significantly lower risk of any STIs (adjusted hazard ratio (aHR) 0.24, 95% CI 0.12-0.47) compared to periods off PrEP after adjustment for socio-demographic characteristics and reasons for initiating PrEP. However, when stratifying for site and type of infection, the risk of extragenital gonorrhea infection (pharyngeal: aHR 2.08, 95% CI 0.85-5.11; rectal: aHR 1.36, 95% CI 0.54-3.46) and extragenital chlamydial infection (pharyngeal: aHR 3.33, 95% CL 0.54-20.36; rectal: aHR 1.73, 95% CI 0.93-3.24) was greater on PrEP compared to off PrEP although these values did not reach statistical significance.

**Conclusions:** The data suggest entry into PrEP care reduced the overall risk of STIs. Service members engaged in PrEP services also receive more STI prevention counseling, which might contribute to decreases in STI risk while on PrEP.

## INTRODUCTION

Consistent use of HIV preexposure prophylaxis (PrEP) reduces sexual risk of HIV acquisition by at least 75% among men-who-have-sex-with-men (MSM).^1^ Since its first approval for HIV prevention in 2012 by the United States (U.S.) Food and Drug Administration (FDA), tenofovir disoproxil fumarate/emtricitabine (TDF/FTC) prescriptions among eligible persons increased from 3% in 2015 to 25% in 2020.^2^ In an effort to increase uptake, federal initiatives in 2019 led to PrEP becoming available free of cost to anyone at high risk for HIV regardless of income or insurance coverage.^2,3^ Although HIV incidence in the U.S. decreased by 8% from 2015 to 2019, reportable sexually transmitted infections (STIs) such as chlamydia, gonorrhea, and syphilis increased by 30% in the same period.^4,5^ Several studies have noted an increase in sexual behaviors that increase the risk of STIs following PrEP initiation including higher rates of sexual partners, condomless anal sex, and sexualized drug use.^6-8^ Reported changes in STI incidence rates and prevalence proportions in MSM PrEP populations have been variable. Some studies demonstrated an association between PrEP use and STI rates following PrEP initiation, although to varying degrees.^6-12^ It is difficult to determine if the increased number of STI diagnoses is consequent to increased frequency of STI testing, as endorsed in the Centers for Disease Control PrEP guidelines.^13^ In contrast, a few studies have shown stable rates of STIs following PrEP initiation, suggesting that the impact of PrEP on risk behaviors and STI incidence is heterogenous in different populations.

The Department of Defense (DoD) Military Health System (MHS) provides health services to 9.6 million beneficiaries. Early adoption of HIV PrEP services within the MHS was driven by infectious diseases subspecialty services located at major military medical treatment facilities (MTFs). However, from 2015-2019, PrEP services slowly expanded to primary care services. Defense Health Agency instruction now mandates that all MTFs have pathways in place to access PrEP services, and that PrEP remain available in both the primary care and infectious diseases subspecialty care settings across the MHS.^14^ The established indications for initiation of PrEP, required laboratory testing, monitoring, and prescribing of PrEP are all conducted in accordance with the current CDC guidelines.^13^ To date, no study has assessed the impact of PrEP initiation on STI incidence in the MHS. Our objective was to assess whether incidence of STIs increased among military service members who were prescribed HIV PrEP.

## METHODS

### Patient Population and Data Collection

We identified service members without HIV who were prescribed the medication TDF/FTC from February 1, 2014 through June 10, 2016 using electronic pharmacy dispensation records. Demographic, service, and sexual risk behavior characteristics of this cohort was described previously.^15^ Health care providers reviewed clinical intake notes in electronic health records (EHRs) to verify initiation of PrEP and extracted reasons for initiation such as men-who-have-sex-with-men (MSM) sexual behavior, lack of condom use, and whether sexual partners were infected with HIV.^15^

For this analysis, two health care providers independently reviewed EHRs and extracted all available positive laboratory results until April 30, 2017 for incident *Chlamydia trachomatis* (CT), *Neisseria gonorrhea* (NG), HIV, hepatitis C virus (HCV), and syphilis prior to PrEP initiation. Longitudinal demographic, laboratory, and pharmacy surveillance records were obtained from the Armed Forces Health Surveillance Division’s Defense Medical Surveillance System (DMSS) from a month before PrEP initiation until March 20, 2019.^16^ Incident cases of STIs starting in the period 30 days prior to, and after initiation of HIV PrEP until March 20, 2019 were assessed from DMSS laboratory records.

### Case definition

A new positive laboratory result for CT or NG was defined as occurring at least 30 days or more after a previous positive result for the same pathogen irrespective of site of collection (for example urine/urethra, pharynx, rectum, vagina, endocervix) or after a previous negative result or, if there were no previous tests, the first positive laboratory test result. A positive laboratory result at any site of collection for the same pathogen on the same day was considered a single infection. History of syphilis before PrEP initiation was defined as the first positive rapid plasma regain (RPR) and *Treponema pallidum* antibody associated with clinical evidence of treatment. After initiation of PrEP, incident syphilis was defined as the first positive RPR test result with a titer of 1:1 or greater followed by evidence of a four-fold rise in titer and a *T. pallidum* positive antibody result. If an RPR titer was missing, incident syphilis was defined as a reactive RPR and a positive *Treponema pallidum* antibody following a nonreactive RPR result within the previous 12 months. Any CT or NG diagnosis ≤30 days following PrEP initiation was considered to have occurred before starting PrEP. An incident STI infection was defined as any new positive laboratory result for CT and NG, syphilis, HCV, and HIV. Service members with no record of a positive laboratory result for STIs were assumed to be free of an STI of interest provided they received care in the MHS.

### Data Analysis

Analysis was restricted to service members with pharmacy dispensation records indicating initiation of PrEP. The frequencies of incident CT, NG, HCV, and syphilis per 100 person-years were calculated for periods on HIV PrEP and off HIV PrEP in the surveillance period. Periods off HIV PrEP included the intervals prior to HIV PrEP initiation, during medication gaps (considered greater than 14 days in duration) and after stopping HIV PrEP until the end of the surveillance period. Follow-up time prior to PrEP initiation was the interval between the later date of start of electronic health records (January 1, 2004) or start date of the first duty assignment in military service, until the date of PrEP initiation. Follow-up time on PrEP was the earliest date of TDF/FTC dispensation until the last day of pill coverage for PrEP; medication gap interval(s) greater than 14 days in duration were deducted from total pill count coverage. Follow-up time after PrEP was calculated from the day pill count ended to the earliest of either end of the study (March 20, 2019), date of separation from service or date of HIV diagnosis.

We fit Andersen-Gill (AG) recurrent event proportional hazards regression models to the data for estimation of unadjusted and adjusted hazard ratios (HRs) and 95% confidence intervals (CIs) of covariates. The counting process time interval proposed by Andersen and Gill is similar to the Cox proportional hazard regression model but allows for subjects to have non-terminal or repeated and multiple events (i.e. occurrence of recurrent or different STIs in the same patient).^17^ The primary covariates of interest in the AG models were HIV PrEP treatment, site and type of STI, and the interaction between site and type of STI and treatment. Other covariates placed additively in the model included mediator variables at PrEP initiation: age, race/ethnicity, marital status, branch of service, pay grade, education attained, occupation, and reasons for starting PrEP. Robust variances based on sandwich estimator were computed in the AG models to adjust for correlation in repeated observations over time in a service member. Characteristics with contiguous levels having similar hazard ratios (all mediator variables except reasons for PrEP initiation) were collapsed in multivariate analyses.

Multicollinearity among covariates in the multivariate model was assessed using variance inflation factor (VIF) and tolerance. Data management and statistical analyses were conducted using Statistical Analysis Software (SAS version 9.4, Cary, North Carolina, U.S) and R Studio (version 4.0.3, Boston, Massachusetts, U.S.).

### Patient Consent Statement

The Walter Reed Army Institute of Research Division of Human Subject Protection (#1861F, #1861G) and the U.S. Army Public Health Center (#146-12.M3) determined the project was a public health activity and not research.

## RESULTS

A preliminary review of health records for 1,700 service members first prescribed TDF/FTC from February 1, 2014, through June 10, 2016 and who were HIV-uninfected at that time revealed that 769 service members initiated PrEP. All 10 female service members were excluded as were 4 others; one person had no evidence of pill dispensation in pharmacy records, an additional two individuals received care entirely in the civilian community, and a fourth service member, who contracted HIV in the surveillance period, was excluded due to self-reported non-initiation of PrEP treatment in an EHR note.

The final analysis included 755 male service members with a total 6340.4 person-years (mean 8.40, range 1.1-15.5) follow-up in the surveillance period from January 1, 2004, to March 20, 2019. Forty (5%) service members had only one visit, 116 (15%) had medication gaps (median 133.0 days, interquartile range (IQR) 66.0-243.0 days) whereas 639 (85%) remained on PrEP continuously. Service members utilized PrEP for a median 635 days (IQR 223-1089) with a median 6 (IQR 3-11) laboratory testing events and a median 77 days (IQR 30-106) between testing events; comparatively, during periods of medication gaps, service members had a median 2 (IQR 2-4) laboratory testing events and a median 83.0 days (IQR 26.0-163.0) between these events.

Overall, 477 (63%) service members had incident STIs (median 2.0, IQR 1.0-4.0), with 151 (20%) identified with only one infection and 326 (43%) having more than one STI (median 3.0, IQR 2.0-5.0); 278 (37%) had no STIs during follow-up. The overall STI incidence in the surveillance period was 21.4 per 100 person-years with a higher incidence during PrEP versus periods off PrEP (38.3 versus 14.4 per 100 person-years, Table 1). The most common STI while off PrEP was from NG (n=288, 5.4 per 100 person-years), which was predominantly urogenital (4.0 per 100 person-years). *Chlamydia trachomatis* infection (n=245, 17.4 per 100 person-years), especially of the rectum (10.1 per 100 person-years), was the most commonly diagnosed STI while on PrEP (Table 1). One service member was diagnosed with HIV while on PrEP and 9 acquired HIV infection an average 16.3 months (range 2.7 to 24.6) after stopping PrEP.

**Table 1.** Incidence of sexually transmitted infections (STI) among 755 male service members from initiation of HIV preexposure prophylaxis, February 1, 2014 - June 10, 2016, through end of follow up.

|  | Not on TDF/FTC* |  | On TDF/FTC^ |  |
| --- | --- | --- | --- | --- |
|  | No. of Infections | Incidence Rate per 100 person-years | No. of Infections | Incidence Rate per 100 person-years |
| Any STI | 714 | 14.46 | 538 | 38.32 |
| *N. gonorrhea* | 288 | 5.83 | 203 | 14.46 |
| Urogenital | 202 | 4.09 | 65 | 4.63 |
| Pharyngeal | 32 | 0.65 | 68 | 4.84 |
| Rectal | 43 | 0.87 | 64 | 4.56 |
| Other | 11 | 0.22 | 6 | 0.43 |
| *C. trachomatis* | 254 | 5.15 | 245 | 17.45 |
| Urogenital | 164 | 3.32 | 83 | 5.91 |
| Pharyngeal | 7 | 0.14 | 17 | 1.21 |
| Rectal | 73 | 1.48 | 142 | 10.11 |
| Other | 10 | 0.20 | 3 | 0.21 |
| *T. pallidum* | 148 | 3.00 | 84 | 5.98 |
| Hepatitis C virus | 15 | 0.30 | 5 | 0.36 |
| HIV | 9 | 0.18 | 1 | 0.07 |

\*Includes events ≤30 days after PrEP initiation, total person-time=4936.578 person-years (gap person-time=577.1498973 person-years, post-PrEP follow-up person-time=82.87200548 person-years)
^Excludes infections diagnosed for test of cure and during TDF/FTC medication gaps; total person-time=1403.877 person-years

Most service members starting PrEP were aged 18-28 years (59%, mean 28.8, range 19.0-56.0), white (47%), and single (72%) (Table 2). The most common reasons for starting PrEP were MSM sexual contact (89%) and/or no condom use (73%). In univariate analysis, service members had a 11% lower risk of incident STIs while on PrEP compared to periods off PrEP (hazard ratio (HR) 0.89, 95% CI 0.73-1.08) although it was not statistically significant (p=0.24)(Table 2). However, in multivariate analysis, risk of an STI lowered significantly by an additional 65% (adjusted HR (aHR) 0.24, 95% CI 0.12-0.47, p <.0001) compared to periods off PrEP after adjustment for age, education level, branch of service, pay grade, occupation, reasons for starting PrEP (MSM/bisexual contact, unprotected sex), and the effect of PrEP on site and type of infection (Table 2). Younger age (18-28 vs 29+: aHR 2.49, 95% CI 1.53-4.04, p<.0001), lower pay grade (E1-E4 vs E5+: aHR 3.09, 95% CI 1.93-4.95, p<.0001), and lack of condom use for starting PrEP (vs not indicated in charts: aHR 1.87, 95% CI 1.15-3.04, p=0.002) were independently associated with STI risk (Table 2).

**Table 2.**
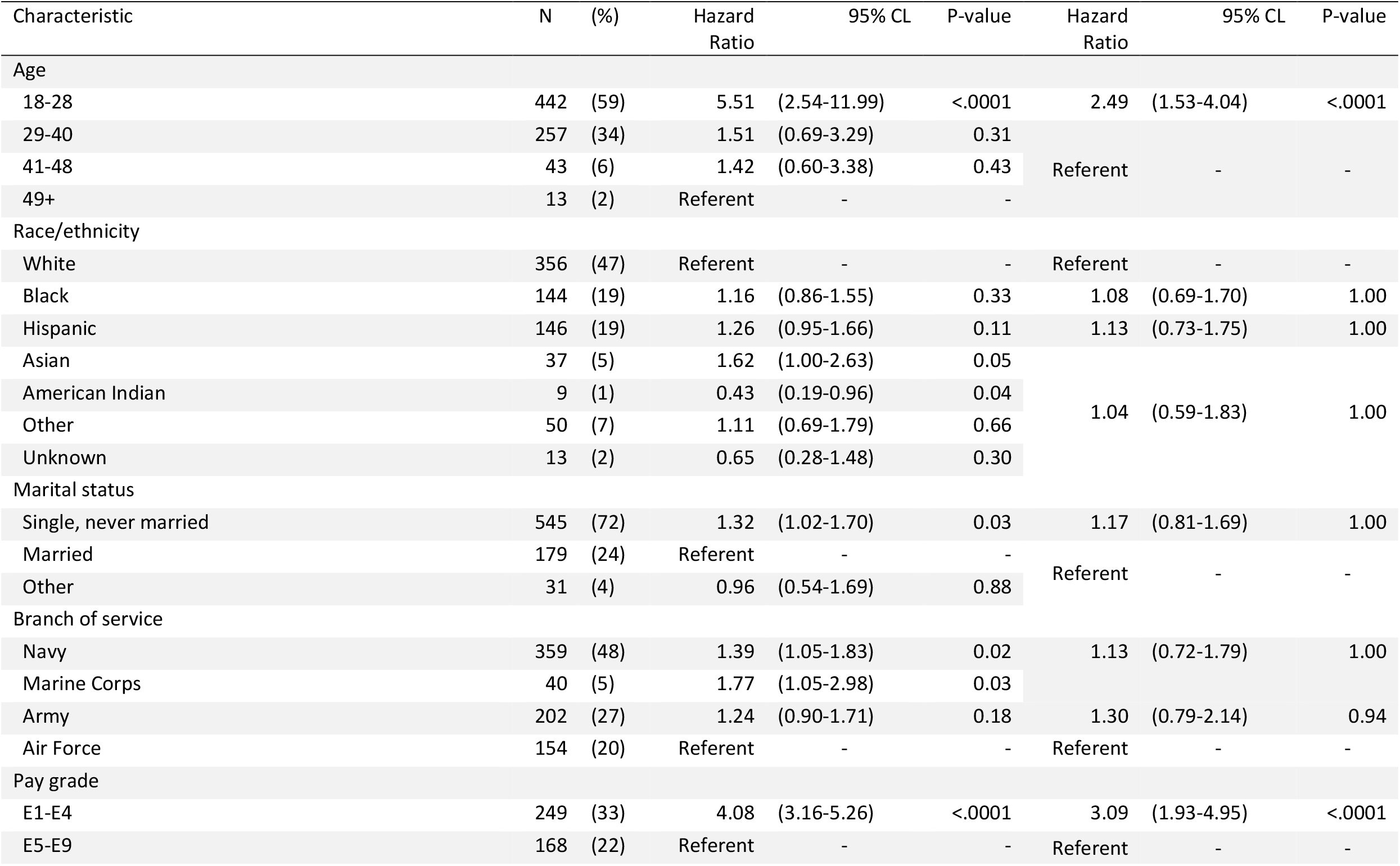
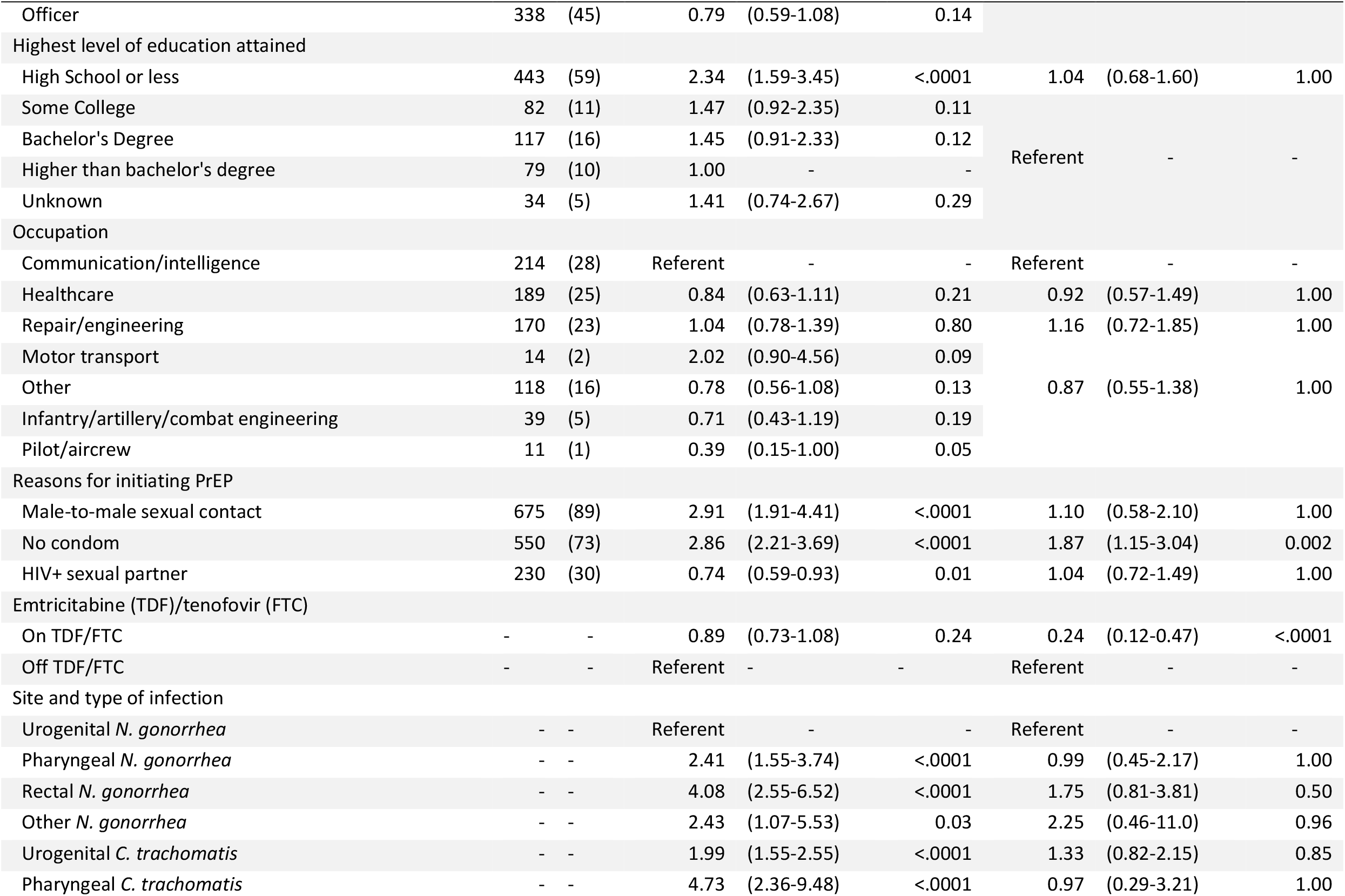
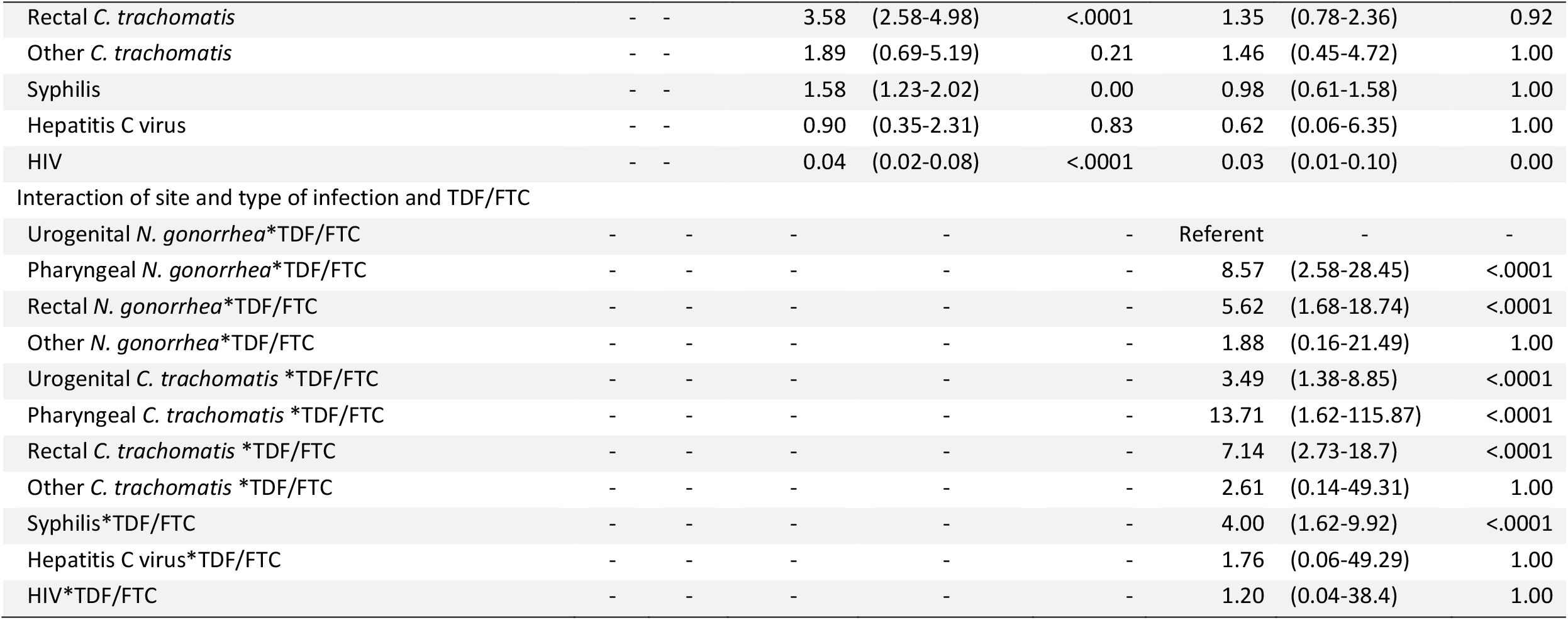
Characteristics of 755 male service members at initiation of HIV preexposure prophylaxis, February 1, 2014 – June 10, 2016, and unadjusted and adjusted hazard ratios of these characteristics associated with incident sexually transmitted infections (STIs) during follow up.

In stratifying the risk of STI by site and type of infection and PrEP use, the risk of diagnosis of urogenital *N. gonorrhea* was significantly lower during PrEP (aHR 0.24, 95% CI 0.13-0.46, p<.0001) in contrast to the risk of STI to periods off PrEP (Table 3). In contrast, the risk of diagnosis of extragenital NG infection (pharyngeal NG: aHR 2.08, 95% CI 0.85-5.11, p=0.21; rectal NG: aHR 1.36, 95% CI 0.54,3.46, p=0.99) and extragenital CT infection (pharyngeal CT: aHR 3.33, 95% CI 0.54-20.36, p=0.49; rectal CT: aHR 1.73, 95% CI 0.93-3.24, p=0.13) was greater on PrEP compared to off PrEP although it did not reach statistical significance (Table 3).

**Table 3.**
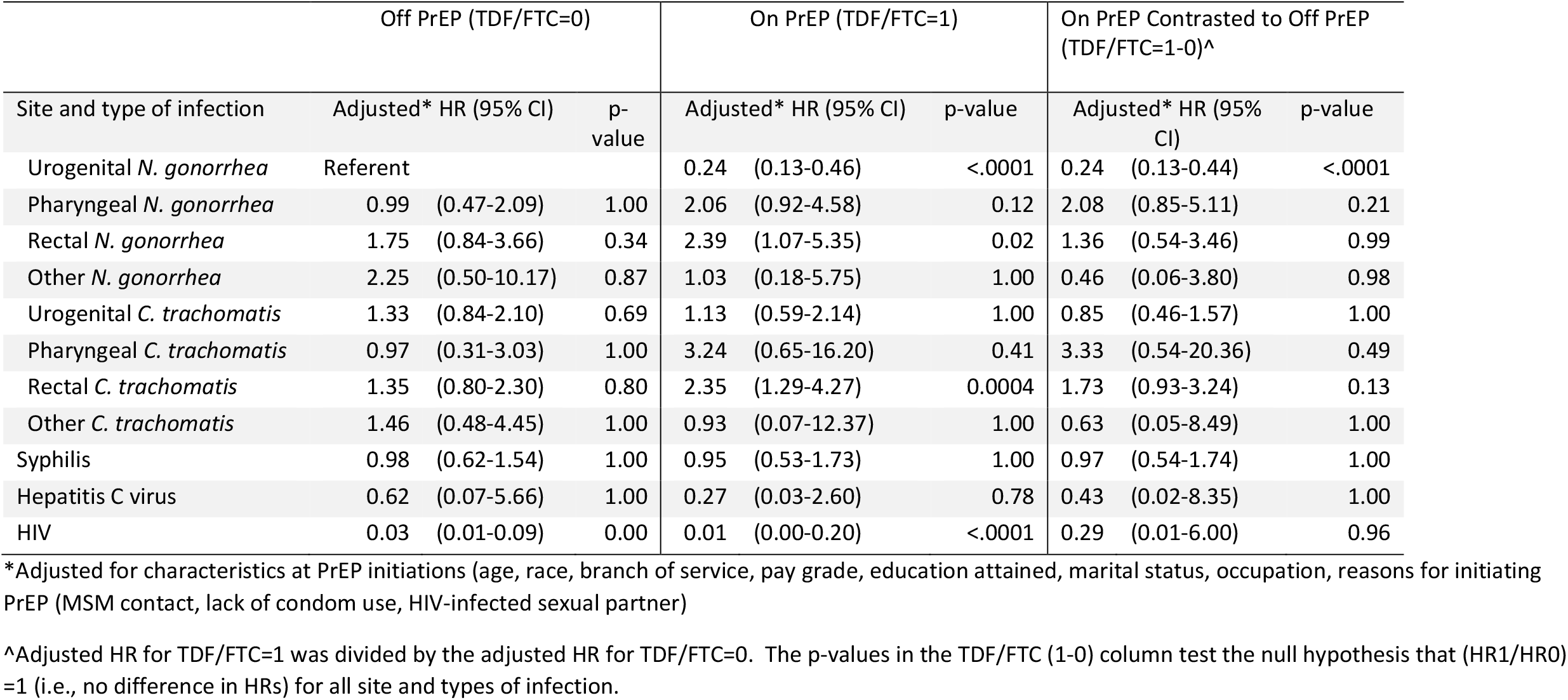
Adjusted hazard ratio (HR) of incident sexually transmitted infections (STIs) by site and type of STI and use of HIV pre-exposure prophylaxis (PrEP) medication (TDF/FTC).

## DISCUSSION

Among male service members who initiated PrEP in the MHS from February 1, 2014 to June 10, 2016, overall STI risk decreased significantly while on PrEP after adjustment for the effect of PrEP on site and type of infection, socio-demographic characteristics and reasons for starting PrEP. However, stratification of risk by site and type of infection and treatment indicated the risk of diagnosis of extragenital bacterial STIs was much greater while on PrEP vs off PrEP although statistical significance was not reached. Younger age (<28 years), lower pay grade (E1-E4), and lack of condom use reasons for starting PrEP were independently associated with STI risk.

STI screening is likely to occur more frequently in the setting of PrEP adherence and follow-up care, in keeping with CDC PrEP guidelines, than off PrEP thus allowing for more opportunities to diagnose asymptomatic infections compared to periods off of PreP. Additionally, testing or screening of extragenital sites is more likely to occur, leading to higher rates of diagnosis of pharyngeal/rectal CT and/or NG. Moreover, service members engaged in PrEP services also receive more STI prevention counseling, which might contribute to decreases in STI risk while on PrEP. These data also suggest service members who were at high risk of STIs appropriately sought PrEP care and were relatively early adopters of prophylactic treatment.

Evidence to date for increased STI acquisition among PrEP users has been mixed. STI positivity did not differ between treatment and control groups nor was an increase in STIs observed in the PrEP group in efficacy trials conducted before July 5, 2020.^1^ These findings may have been confounded by study design and inclusion criteria since patients enrolled in trials were at high risk of HIV infection and blinding to PrEP allocation and whether treatment was received may have influenced sexual behavior. However, an increased odds of overall STI diagnosis was observed (odds ratio (OR), 1.24, 95% CI, .99-1.54) in a meta-analyses of open-label PrEP studies (2014-2017), although it was not statistically significant despite significantly increased odds of rectal STIs (OR, 1.39; 95% CI, 1.03–1.87), especially for chlamydia (OR 1.59, 95% CI 1.19-2.13).^18^ Although these studies were limited by lack of pre-enrollment analysis of STI positivity since studies compared STI positivity at baseline to follow-up. In studies which did address STI history before PrEP was initiated, an increase in incidence (Nguyen et al. adjusted rate ratio (aRR) 1.39, 95% CI 0.98–1.96; Beymer et al. RR 1.36, 95% CI 1.06–1.74; Traeger et al. aRR 1.21, 95% CI 1.06-1.39) was observed although statistical significance was not reached in all analyses. These cohort studies compared STI diagnosis among MSM in sexual health care in up to 12 months preceding PrEP initiation to follow-up.^12,19,20^ While our analysis demonstrated an increase in crude incidence during periods on PrEP versus off PrEP, by stratification for site and type of infection, we were able to show that overall STI risk decreased, contrasting with trends from other cohort studies. This further highlights the non-uniform impact that PrEP initiation has on non-HIV STIs.

In our analysis, despite the independent association between STI risk and lack of condom use at baseline, STI incidence (38.3 per 100 person-years) was much lower compared to reports of similar cohort analyses (83.5-107.7 per 100 person-years).^12,19,20^ It is possible the difference may be due to shorter follow-up time in these studies and/or differences in STI prevalence across geographically different sexual networks since level of condom use was not associated with STI risk in one of the studies.^12^

The higher STI risk seen among younger male service members in our analysis is reflective of increasing STI incidence over time, in younger age groups both in the military and the general population irrespective of PrEP use.^21,22^ In fact STIs have posed a medical readiness issue historically in the 20^th^ century to U.S. and armed forces worldwide, especially among younger enlisted personnel.^23^ Even recently an analysis of U.S. military EHRs from 2012-2020 showed STI incidence was highest among <29-year-old age group and enlisted personnel, especially those in junior ranks (EI-E4).^21^ In the U.S. general population, the 15–24-year-old age group accounts for at least half of all reportable STIs with younger MSM (18-29 years) at highest risk of bacterial STIs, especially syphilis.^24-26^ Adolescents and young adults are thought to be at higher risk than adults due to social, behavioral, and biological factors such as lower utilization of sexual health services especially in the southern U.S., biological susceptibility among younger females, childhood adversity, and concurrent partners.^27^

Our analysis has several limitations. Differential screening for STIs while in PrEP care to periods off PrEP may have introduced surveillance bias and differential detection of STIs during follow-up. However, a strength of this analysis was that surveillance for any prior STI diagnosis from entry into service to initiation of PrEP care served as a control period; this may have minimized large differences compared to surveillance that was limited to only 3 to 12 months before PrEP initiation. Second, since pharmacy dispensation records were used to assess PrEP coverage and not self-reported pill counts or TDF/FTC blood levels at each visit, follow-up time in PrEP care could have been misclassified leading to error in estimations of risk. Third, the case definition of syphilis changed from before PrEP initiation to follow-up. This may have led to error in estimation of syphilis risk and, consequently, STI risk.

In conclusion, although uptake of PrEP care may have led to a modest increase in diagnosis of STIs, especially of extragenital infections, the data suggest entry into PrEP care reduced the overall risk of STI. This is likely multifactorial: service members who sought PrEP may have recognized their existing high risk of HIV acquisition and adopted PrEP early and they may have benefited from greater STI prevention counseling. Nonetheless, the data suggest routine STI screening should remain a cornerstone of PrEP care given the STI risk in this population.

## Data Availability

For ethical reasons which include possible adverse impact to employment of personnel still in military service, data cannot be made publicly available. For questions about the data, please contact the WRAIR Public Affairs Office at

## Acknowledgements

We are grateful to Christine Walsh for efforts in case validation; Leslie Clark and Angelia Eick-Cost for facilitating data requests from the Armed Forces Health Surveillance Division (Silver Spring, Maryland).

## Financial Support

This study was supported in part by the U.S. Army Medical Research and Development Command under Contract No W81XWH-16-C-0225 and by the Combined HIV and Infectious Disease Agreement (CHIDA) cooperative agreement (W81XWH-18-2-0040) between the Henry M. Jackson Foundation for the Advancement of Military Medicine, Inc. (HJF), and the U.S. Department of Defense.

## Conflicts of Interest

The authors have no conflicts of interest to declare.

## Notes

### Competing Interest Statement

The authors have declared no competing interest.

### Author Declarations

xI have followed all appropriate research reporting guidelines and uploaded the relevant EQUATOR Network research reporting checklist(s) and other pertinent material as supplementary files, if applicable.

